# Using a mixed-reality headset to elicit and track clinically relevant movement in the clinic

**DOI:** 10.1101/2024.07.07.24310049

**Authors:** Dylan Calame, Evan Lester, Phil Chiu, Lauren Seeberger

## Abstract

**Background:** 21st century neurology will require scalable and quantitative tools that can improve neurologic evaluations over telehealth and expand access to care. Commercially available mixed-reality headsets allow for simultaneous presentation of stimuli via holograms projected into the real world and objective and quantitative measurement of hand movement, eye movement, and phonation.

**Methods:** We created 6 tasks designed to mimic standard neurologic assessments and administered them to a single participant via the Microsoft HoloLens 2 mixed-reality headset. The tasks assessed postural hand tremor, finger tapping, pronation and supination of hands, hand and eye tracking of a center-out task, hand and eye tracking of a random motion task, and vocal assessment.

**Findings:** We show the utility of the HoloLens for commonly used neurological exams. First, we demonstrate that headset-derived holograms can project hand movements and objects in 3D space, providing a method to accurately and reproducibly present test stimuli to reduce test-test variability. Second, we found that participant hand movements closely matched holographic stimuli using a variety of metrics calculated on recorded movement data. Third, we showed that the HoloLens can record and playback exam tasks for visual inspection, sharing with other medical providers, and future analysis. Fourth, we showed that vocal recordings and analysis could be used to profile vocal characteristics over time. Together, this demonstrates the versatility of mixed reality headsets and possible applications for neurological assessment.

**Interpretation:** Administering components of the neurologic exam via a self-contained and commercially available mixed-reality headset has numerous benefits including detailed kinematic quantification, reproducible stimuli presentation from test to test, and can be self-administered expanding access to neurological care and saving hospital time and money.

**Funding:** This work was supported by grants from the National Institutes of Health (NIH) (F30AG063468) (E.L.), (F31NS113395) (D.J.C), and the Pilot Grant Award from the University of Colorado Movement Disorders Center (D.J.C).

## Introduction

Characterization of movement is a critical aspect of the neurological examination used to assess motor pathology. Movements of the eyes, movements of the limbs, gait, posture, and phonation are commonly assessed to detail the severity and progression of disease. Neurologists assess these characteristics using agreed upon scales for different disease classes. Clinicians rate various features of movement and behaviors pathognomonic for that disease using an integer scale. For cerebellar ataxia, clinicians use the Scale for the Assessment and Rating of Ataxia (SARA), for Parkinson’s disease the Unified Parkinson’s Disease Rating Scale (UPDRS), and for Huntington’s disease the Unified Huntington’s Disease Rating Scale (UHDRS)^1–3^.

While studies point to consistency of the inter-rater reliability between neurologists scoring motor pathology according to these scales^4^, select aspects of these exams have lower reliability, bringing into question the usefulness of rating these movement features for classifying disease severity^5^. Often, these difficult-to-score components of the exam involve the clinician attempting to assess features of fast movements, like in the finger chase and fast alternating hand movements tests in the SARA. Introducing objective quantification of movement using motion capture technology promises improvement in disease severity assessments, however the hardware and software necessary for this type of motion capture is often cumbersome, difficult to use, and costly in terms of time and resources. Further, variation in the technique of the healthcare professional administering the exam can lead to differences in the movements evoked, obscuring the results of exams performed across providers or medical sites. Thus, assuring that the exam is performed the same way is critical to minimizing test-retest variability.

A novel way to both present stimuli in a consistent manner and to record behavioral responses is with mixed reality headsets. The Microsoft HoloLens 2 is a commercially available headset that is worn like a pair of sunglasses. It renders 3D holograms of objects visible to the user, superimposed on the real world. The user is then able to interact with holograms in this semi-virtual environment (Figure 1 shows an example of a holographic sphere superimposed upon a brick wall in the real world). In addition to holographic rendering, the HoloLens 2 has built in eye tracking, fully articulated camera-based hand tracking, and a microphone for vocal analysis. Unlike fully-immersive virtual reality devices, which are associated with motion sickness in some users^6–8^, mixed reality is not fully immersive and causes less visual-vestibular conflict^9^, making it a suitable candidate for clinical applications. These capabilities make it an ideal device to both guide patients with movement disorders through a standardized neurological physical exam and provide clinicians and researchers with high-fidelity behavioral characterizations.

**Figure 1.**
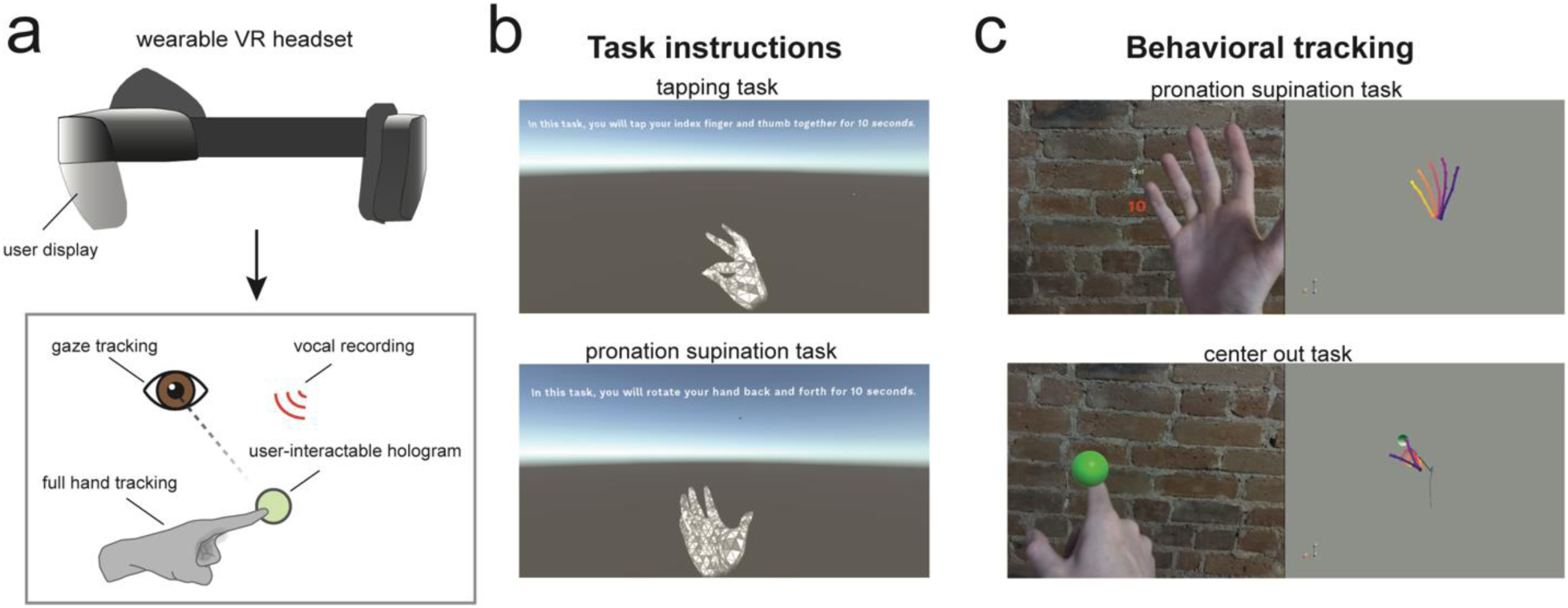
Using the Microsoft HoloLens 2 mixed-reality headset to elicit and track clinically meaningful behavior. **a.** To elicit and track clinically relevant behavior, users wear a mixed-reality headset (top graphic). This headset can display holograms to users while tracking the position of 26 joints on the user’s hand, the user’s gaze, and the user’s voice. **b.** For each task, users are displayed a set of written instructions. The tapping task and the pronation-supination task also have holographic hands that visually guide users through the task. **c.** Display from an egocentric vantage point of the user’s experience (left) compared to the tracked behavioral data analyzed offline (right). The egocentric view in the top panel shows the countdown displayed to the user during the pronation-supination task and on the bottom shows the user interacting with a green sphere hologram in the center out task.

Here, we designed a suite of virtual tasks to elicit behaviors of interest in a neurological examination. Tasks are designed to be self-paced by the user and fully administered by the headset while providing robust behavioral data for the purposes of neurological assessment. This approach has several benefits. First, it can free up the time of medical staff as data can be collected in an asynchronous manner that does not require the physical presence of a highly trained physician. Second, it can administer perfectly reproducible stimuli and provide high-fidelity measurements. This could improve quantification of neurological changes over time and in response to medications or interventions. Third, it democratizes access to quality assessment of neurological status. Many patients, especially in rural areas, do not have access to trained movement disorder staff. A commercially available and mass-produced headset like the HoloLens 2 theoretically allows patients to wear the headset at home or in a primary care setting and upload their results to a qualified physician for interpretation. These results could be followed up via telehealth appointments to discuss results. Fourth, presentation of holographic stimuli using the HoloLens 2 allows for quantification of novel tasks that are not possible in a standard neurological examination. This can allow for the discovery of new and clinically relevant psychophysical metrics of movement disorders that have yet to be described. All data displayed in this study are from a single member of the study staff to provide proof of concept. Future studies will aim at using this application in a patient population that have movement disorders of interest.

## Results

### Task design and instruction

Task instructions, stimuli, and participant recording were designed and tested in Unity software and uploaded to the HoloLens headset with Visual Studio (Figure 1a). We designed 6 tasks to mimic several commonly used neurological exam maneuvers: **1)** A tremor assessment task where users attempt to hold their hands still in front of them for 10 seconds; **2)** A tapping task where users are instructed to tap their index finger and thumb together as fast as they can, while spreading the thumb and index fingertips as wide as possible on each tap, for 10 seconds; **3)** A pronation-supination task where users are instructed to turn their hand back and forth as fast as possible, while turning as far as possible in each direction, for 10 seconds; **4)** A center out task where users interacted with a holographic sphere that would move in 1 of 8 radial directions and users were instructed to track it with their fingertip; **5)** A random tracking task where users are instructed to track the position of a holographic sphere moving in a pseudo-randomized path with their fingertip; **6)** And a vocal assessment task where users read phrases presented to them on their display aloud.

Each task is introduced to the user using written instructions displayed on the screen of the headset. In the finger tapping and pronation-supination tasks, holographic displays of the hand were presented to the user as a visual guide to supplement the written instructions (Figure 1b). Once the user has read the instructions, they start the task by selecting a start icon with their fingertip or by saying “Start”.

For the tapping, pronation-supination, center out, and random tracking tasks users completed the tasks first with their right hand and then their left. For tasks involving hand movements (tasks 1-5), positional data for was collected for 26 points on each hand and gaze data was collected to assess eye movements at 60 Hz (see Methods). In the vocal tracking task, audio recordings and gaze data were collected.

### Postural tremor assessment task

Tremors are the most common movement disorder and can be classified as resting, postural, or action tremors. Postural tremors can be seen in physiologic tremor, essential tremor, metabolic disturbances, and Parkinson’s disease^10,11^. To test for and quantify postural tremor, users were instructed to hold both hands and arms straight out in front of them for 10 seconds (Supplemental Video 1). Once the headset had detected that the hands are visible and have crossed a positional threshold in the outward direction, it initiated a 10-second countdown displayed to the user. At the end of the 10 seconds users are instructed that they may put their hands back down. This task is presented first so that the length of the user’s arms can be measured during the hold period and can be utilized in later tasks to appropriately place interactable holograms within the user’s reach.

To quantify the characteristics of a user’s postural tremor we isolated the position, speed, and acceleration of the center of the right and left hands during the 10-second countdown (Figure 2a). Measuring the average speed and acceleration magnitude in the tracked point during this window provided a quantitative score reflecting the stillness of the user’s hands (right hand: speed = 0.0147 ± 0.0086 m/s, acceleration = 0.549 ± 0.293 m/s^2^; left hand: speed = 0.0197 ± 0.0131 m/s, acceleration = 0.786 ± 0.486 m/s^2^; mean ± SD).

**Figure 2.**
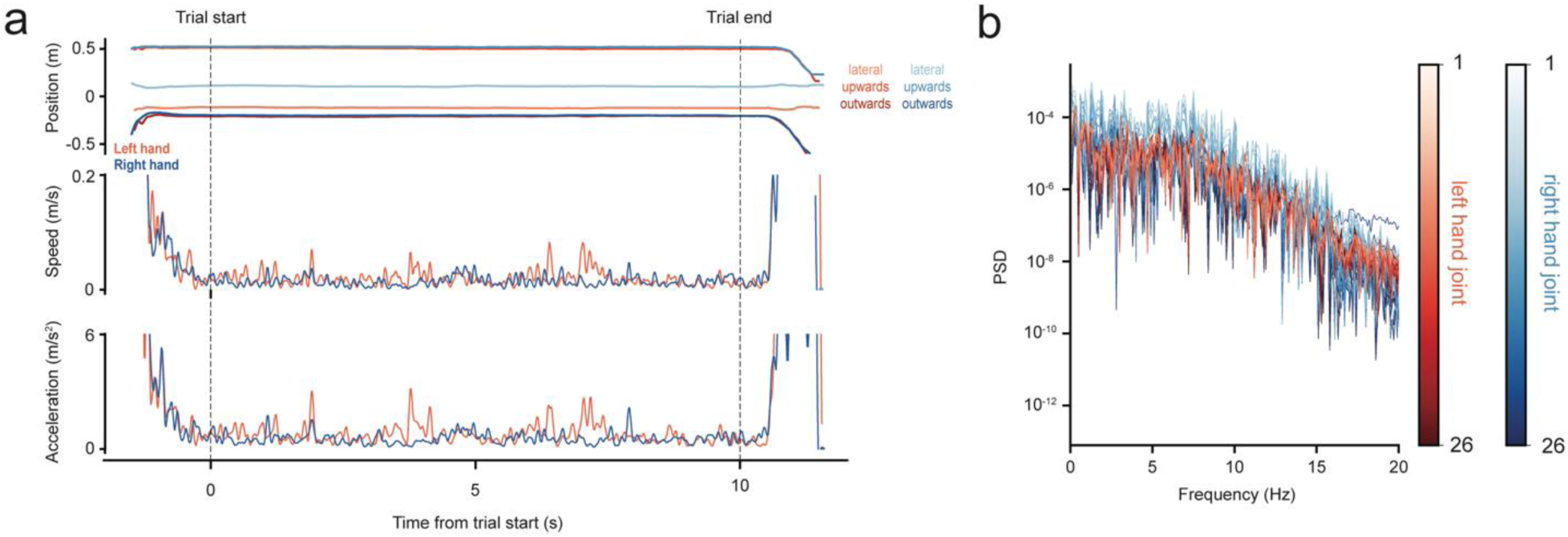
Hand tremor quantification evoked during the tremor assessment task. **a.** Position (top), speed (middle), and acceleration (bottom) traces of the right and left hand. Dashed lines indicate the start and end of the trial countdown. For the positional data, traces are broken down into the lateral, upwards, and outwards directions. **b.** Power spectral density for each tracked joint of the right and left hand.

To further characterize the presence of tremor and tremor amplitude in each tracked point in the hands, we performed a fast Fourier transform of the speed at each of the 26 tracked points of each hand. We assessed the power spectral density (PSD) of each point across the frequencies from 1-20 Hz to identify tremors in each joint (Figure 2b). Patients with tremors exhibit peaks that could be identified in the PSD analysis, with precise understanding of the magnitude of tremor in each of the tracked joints, providing a detailed fingerprint of the idiosyncratic tremor that individual patients might exhibit.

To confirm that the motion captured by the headset was an accurate quantitative description of the movements performed, we had the user simulate a tremor at different frequencies at the rate of an external auditory stimulus delivered by a metronome. The user moved hands rhythmically in the vertical dimension at a rate of 1, 2, 3, and 4 Hz. Analyzing the peak of the PSD of the vertical positional traces for the palm in the right and left hands revealed a strong correlation with the driving frequency of the metronome (Figure 1Sa; Right hand: linear regression of metronome frequency and peak PSD frequency: slope = 0.94, p-value = 2.26 x 10^-4^; Left hand: linear regression of metronome frequency and peak PSD frequency: slope = 0.93, p-value = 3.47 x 10^-4^).

### Tapping task

Disruptions in finger tapping of the thumb and index finger can be seen in Parkinson’s disease^12^, atypical parkinsonism^13^, upper motor neuron demyelinating diseases such as multiple sclerosis^14,15^, and Alzheimer’s disease^16^. Disease specific changes can be seen in the speed, amplitude, and regularity of tapping^16^. To probe finger tapping movements we designed a task that instructed users to tap their thumb and index fingers together for 10 seconds (Supplemental Video 2). The instructions included an animated hologram of a right or left hand tapping to improve the user’s understanding of the task. Once users had time to read the instructions, they were instructed to push a holographic start button to begin the task. Once the task was started, users were instructed to hold their right hand up in front of them. Once the headset detected the user’s hand, it began a 5-second countdown with an instruction for users to begin tapping at the end the countdown. A go-cue at the end of the countdown instructed users to begin tapping at which point they were presented with another 10-second countdown. At the end of the 10 seconds, they were told they were able to put their hand down, and they were presented the same series of instructions for the left hand.

Timestamps indicating the time of the go-cue and end of the countdown were used to isolate the tapping interval from saved positional data for trials of the right and left hand (Figure 3a,e). A tap amplitude timeseries was extracted by taking the Euclidean distance between the tip of the thumb and index finger (Figure 3b,f). Isolating local minima and maxima of this amplitude timeseries allowed us to isolate the start and end of individual taps, which we used to compute the time and amplitude of each tap (Figure 3c,d,h,g; Right hand: tap time = 0.119 ± 0.012 s, tap amplitude = 0.234 ± 0.018 m, n = 42 taps; Left hand: tap time = 0.250 ± 0.018 s, tap amplitude = 0.137 ± 0.007 m, n = 38 taps; mean ± SD). The slope of a linear regression fit to the tap amplitudes and tap times across the trial was used to assess movement decrement (Right hand: linear regression of tap amplitude: slope = -2.48 x 10^-4^, p-value = 0.10; linear regression of tap time: slope = 1.51 x 10^-4^, p-value = 0.53; Left hand: linear regression of tap amplitude: slope = -3.27 x 10^-4^, p-value = 0.001; linear regression of tap time: slope = 3.95 x 10^-6^, p-value = 0.99).

**Figure 3.**
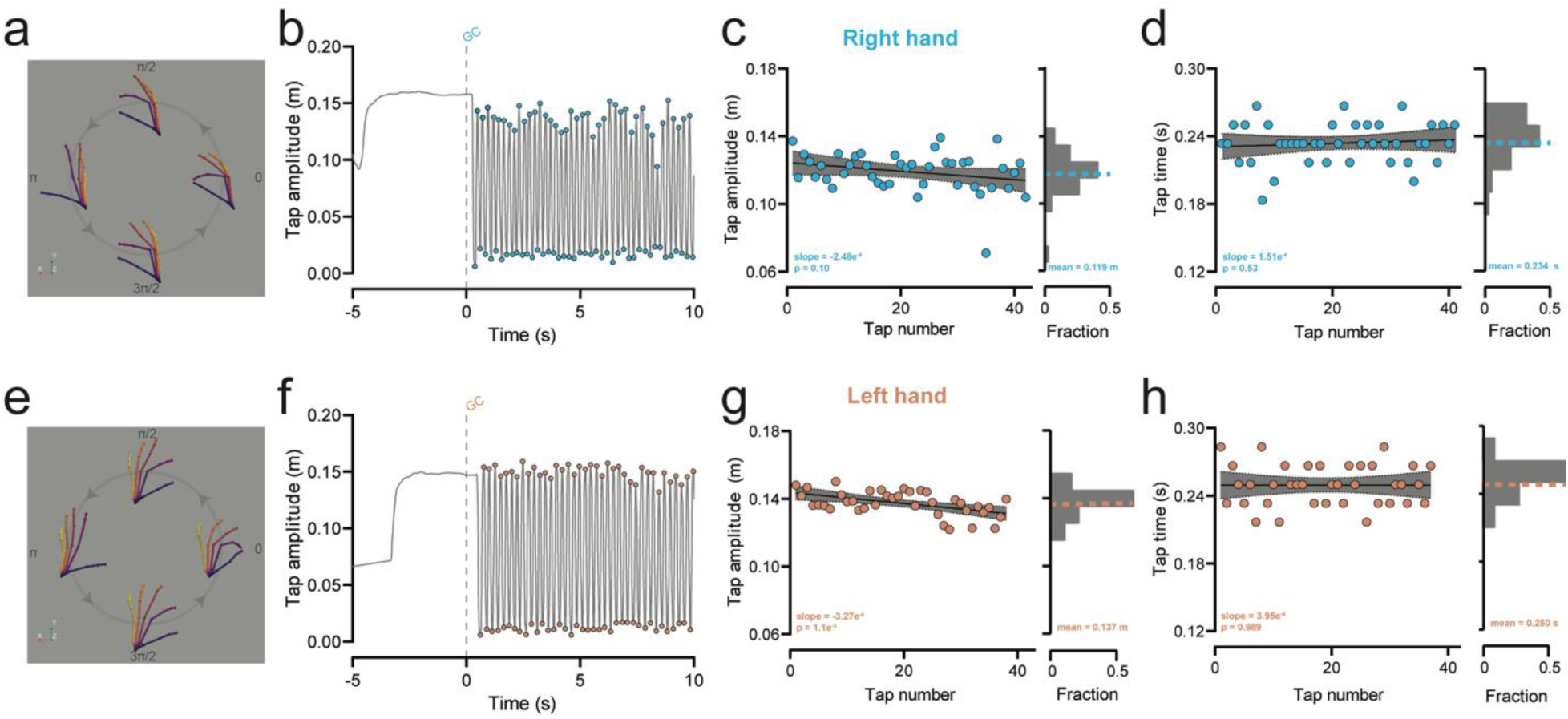
Quantification of hand movement evoked with the tapping task. **a, e.** Representative examples of data hand collected in a single tapping cycle for the right (**a**) and left (**e**) hands. **b, f.** Extracted tap amplitude (Euclidean distance between the thumb tip and index tip) for the right (**b**) and left (**f**) hands. The dashed line indicates the time the go-cue was presented to the user (GC). Colored dots indicated the extracted times of the minima and maxima of each tap. **c, g.** Left: quantification of the tap amplitude for each tap in the trial for the right (**c**) and left (**g**) hand. The line indicates the best fit line of the linear regression with a 95% confidence interval. Right: histogram of tap amplitudes. Mean tap amplitude is shown with a dashed line. **d, h.** Same as the analysis presented in **c** and **g** but instead quantifying tap time.

To assess whether the thumb and finger motions captured reflected the rate of hand movements, we had the user tap at a rate specified with an auditory tone delivered by a metronome across a range of frequencies (80, 120, and 160 bpm). Tap interval time closely matched the metronome beat interval for both the right and left hand (Figure 1Sb; Right hand: linear regression of metronome beat interval time and tap interval time: slope = 1.04, p-value < 1 x 10^-15^; Left hand: linear regression of metronome beat interval time and tap interval time: slope = 0.99, p-value < 1 x 10^-15^).

### Pronation-supination task

Pronation-supination movements are another commonly used exam task in the assessment of Parkinson’s disease^17^. We designed a similar task to finger tapping where users were instructed to rotate their hand back and forth for 10 seconds (Supplemental Video 3). Instructions and task structure followed the format described in the finger tapping task and included a holographic animation to guide the users’ movements.

To quantify pronation-supination, we measured the turn angle around a vector projecting from the wrist to the metacarpophalangeal (MCP) joint of the middle finger (see Methods). Local minima and maxima were calculated from turn angles to extract single turn cycles which we then used to analyze turn angle amplitudes and turn times (Figure 4a-h; Right hand: turn time = 0.376 ± 0.097 s, turn angle amplitude = 187.9 ± 15.9 degrees, n = 24 turns; Left hand: turn time = 0.410 ± 0.033 s, turn angle amplitude = 182.0 ± 7.7 degrees, n = 24 turns; mean ± SD). The slope of a linear regression fit to the turn angle amplitudes and turn times across the trial was used to assess movement decrement (Right hand: linear regression of turn angle amplitude: slope = 0.123, p-value = 0.80; linear regression of turn time: slope = 1.58 x 10^-3^, p-value = 0.55; Left hand: linear regression of turn angle amplitude: slope = -0.348, p-value = 0.13; linear regression of turn time: slope = 2.40 x 10^-3^, p-value = 0.01).

**Figure 4.**
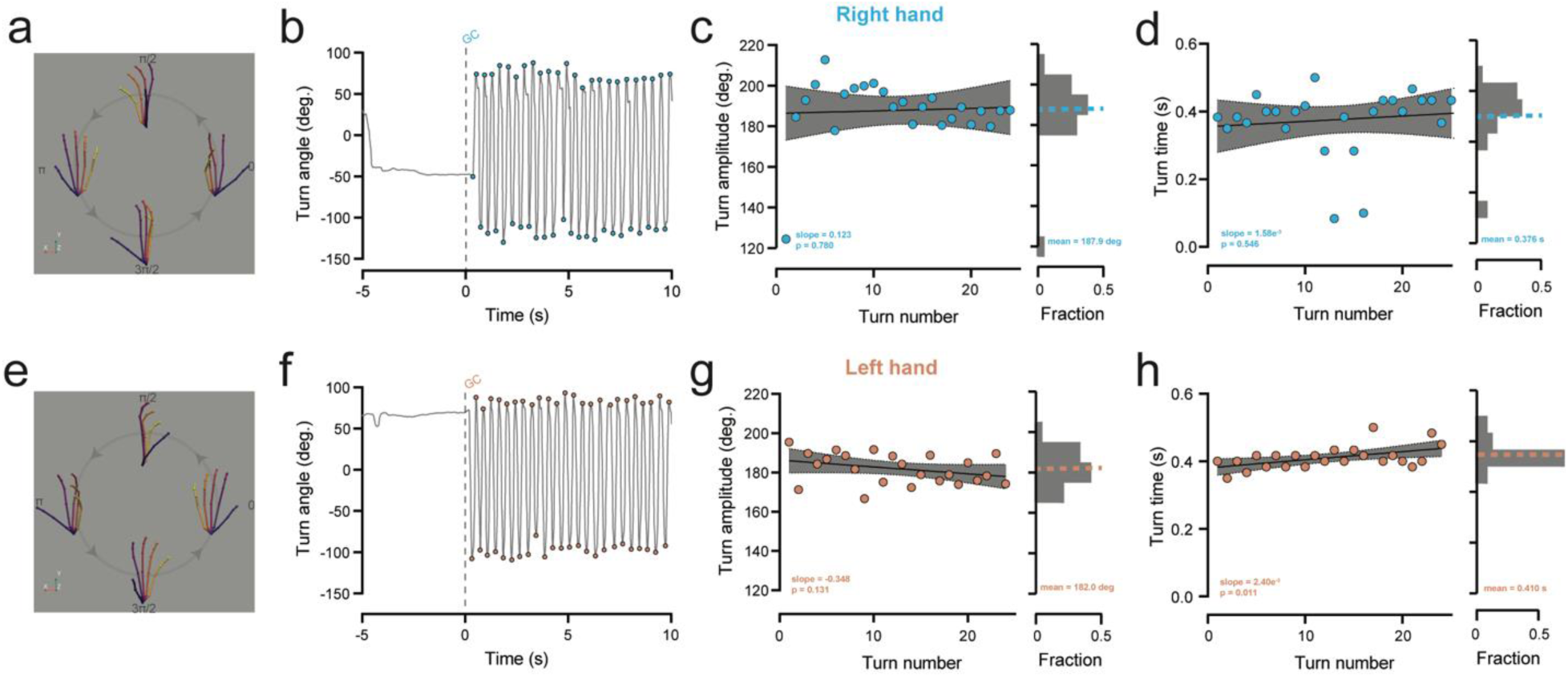
Quantification of hand movement evoked with the pronation-supination task. **a, e.** Representative examples of data hand collected in a single pronation-supination cycle for the right (**a**) and left (**e**) hands. **b, f.** Extracted turn angle for the right (**b**) and left (**f**) hands. The dashed line indicates the time the go-cue was presented to the user (GC). Colored dots indicated the extracted times of the minima and maxima of each turn. **c, g.** Left: quantification of the turn amplitude for each turn in the trial for the right (**c**) and left (**g**) hand. The line indicates the best fit line of the linear regression with a 95% confidence interval. Right: histogram of turn amplitudes. Mean turn amplitude is shown with a dashed line. **d, h.** Same as the analysis presented in **c** and **g** but instead quantifying turn time.

To assess whether captured hand rotations reflected the rate of actual hand pronation-supination movements, we had the user rotate their hand at a rate specified with an auditory tone delivered by a metronome across a range of frequencies (80, 120, and 160 bpm). Turn interval time closely matched the metronome beat interval for both the right and left hand (Figure 1Sc; Right hand: linear regression of metronome beat interval time and turn interval time: slope = 0.85, p-value < 1 x 10^-15^; Left hand: linear regression of metronome beat interval time and turn interval time: slope = 0.85, p-value = 2.61 x 10^-12^).

### Center out task

To assess the users’ ability to track a moving target with their hands and eyes, assess for the presence of an action tremor, and measure a patient’s reaction time, we designed a task where users are instructed to track a moving holographic sphere with their fingertip and eyes as it moves from the center to the periphery and back to the center (Supplemental Video 4). Action tremors can be indicative of cerebellar lesions and help differentiate between Parkinson’s disease and essential tremor^11^ and slowed reaction times can also be indicative of Parkinson’s disease^18^. The target is a 4-cm diameter grey sphere that starts at 75% of the user’s reach established in the tremor assessment task. When the user touches the sphere, it turns green to provide visual feedback during the task. The sphere starts at a central location, and after the user has touched the target for 1 second it moves 20 cm to 1 of 8 eccentric locations. The user is then supposed to move as quickly and accurately to the target as possible such that they touch the target again. After the target has been touched in the eccentric location for 1 second, it moves 20 cm back to the starting central location, and users track it again back to this location. We refer to movements to the eccentric targets as “center-out” trials and movements back to center as “return-to-center” trials (Figure 5a). The task consists of 48 total trials, such that there are 3 center-out movements to each of the 8 eccentric targets, and 3 return-to-center trials from those targets. This task is conceptually similar to the finger chase exam commonly used in the scale for the assessment and rating of ataxia (SARA)^3^.

**Figure 5.**
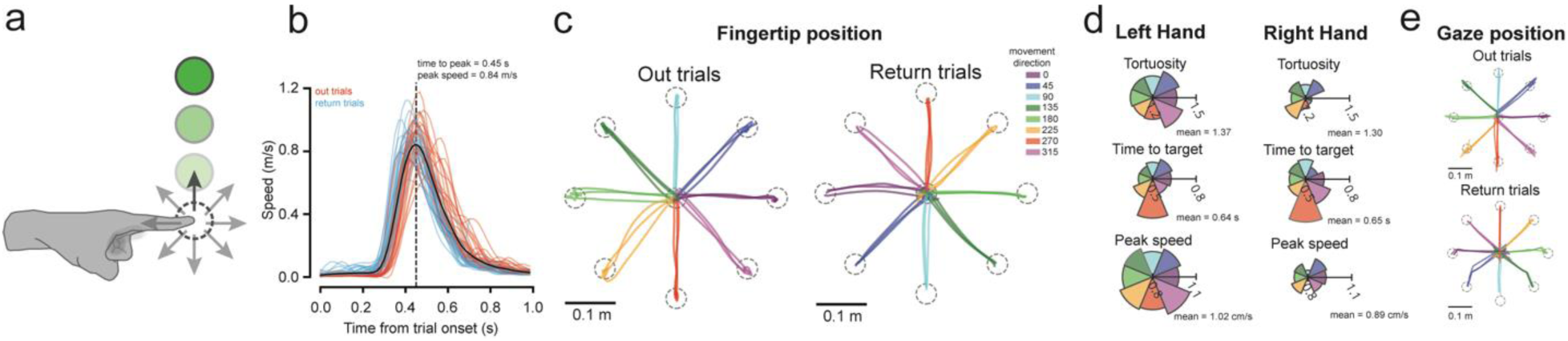
Movement quantification evoked with the center out task. **a.** Users were instructed to touch a holographic sphere with the tip of their index finger. After touching the sphere for 1 second, it would move in 1 of 8 directions and users would attempt to follow its motion with their fingertip. After touching it for 1 second at the new location the target would move back to the origin position. **b.** Speed profiles of the fingertip from the onset of each trial. Center-out trials are shown in red and return-to-center trials are shown in blue. The mean speed profile is shown in black. Time of mean peak speed is shown as a vertical dashed line. **c.** Positional traces of the fingertip in center-out and return trials. Color of the trace indicates the movement direction (in degrees). Dashed circles represent the starting and final positions of the sphere. **d.** Polar plots showing the mean tortuosity (top), time to target (middle), and peak speed (bottom) in each movement direction for the left and right hands. The scale is shown with a horizontal bar in each polar plot. **e.** Positional traces of the gaze point during the trials shown in **c**.

In each trial, times indicating when the target started moving to a new location were used to divide positional traces of fingertip positions into discrete trials. First, we analyzed reaction times by isolating speed profiles aligned to the trial onset time (Figure 5b; Statistics for right hand data shown in figure 5b: peak speed = 0.84 m/s, time to peak speed = 0.45 s). To quantify clinically relevant features of movement we analyzed tortuosity as a proxy for dysmetria and action tremor, time to target as a proxy for accuracy, and peak speed (Figure 5c,d). We defined tortuosity as the total path length of a trial’s positional trajectory divided by the distance between the target start and endpoint. We also calculated the peak speed of each movement. We plot each of these metrics for trials subdivided by the direction of the movement from the start point to the endpoint to display any asymmetrical impairment of movement (Right hand mean of all trials: tortuosity = 1.30, time to target = 0.65 s, peak speed = 0.89 cm/s; Left hand mean of all trials: tortuosity = 1.37, time to target = 0.64 s, peak speed = 1.02 cm/s).

We also tracked eye angle during these trials. To determine the gaze point we found the shortest line between the gaze ray and the target. The location where this line intersected the gaze ray we refer to as the “gaze point”. We use this point to measure the 3D position that the user was visualizing at each timepoint (see Methods). Analysis of gaze point data divided into trials shows that it closely follows the trajectory of the fingertip on individual trials (Figure 5e).

### Random tracking task

Dysmetria is the inability to control the distance, speed, and range of motion necessary to perform smoothly coordinated movements and can be due to cerebellar damage from trauma, tumors, or demyelinating diseases^19^. We designed a random tracking task to assess a user’s ability to follow a holographic target moving in a pseudo-random path that is analogous to the finger-chase and smooth pursuit tasks common to neurologic exams (Figure 6a, Supplemental Video 5). Quantifying these movements is difficult by physicians in a clinical setting, but the high-fidelity behavioral tracking possible with the HoloLens 2 allows for design of tasks that would have previously been impossible and may provide a new and clinically relevant metrics.

**Figure 6.**
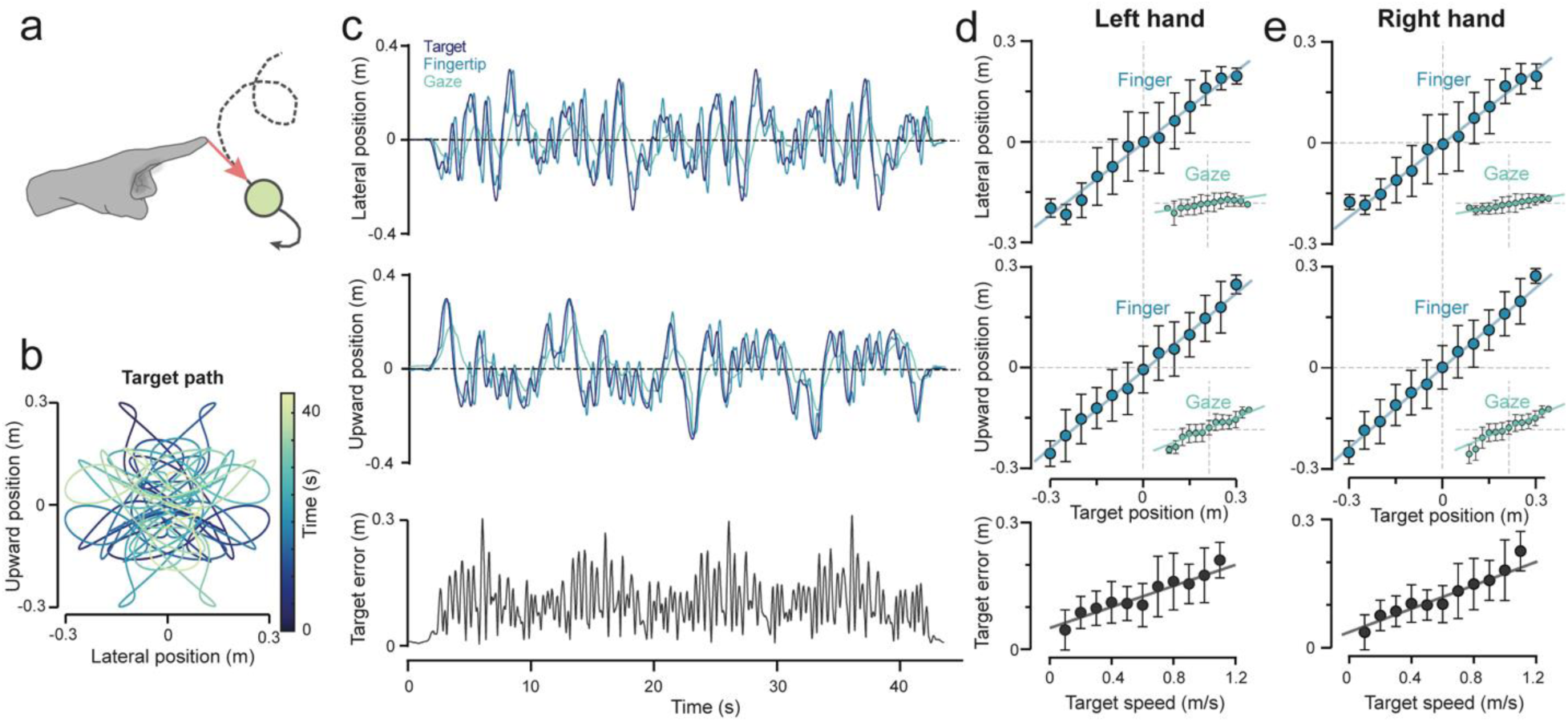
Quantifying eye and hand movements evoked with the random tracking task. **a.** Users were instructed that they would be attempting to track a holographic sphere with their index finger as it traversed the workspace in front of them in a pseudo-random path. **b.** Holographic sphere trajectory in the upward and lateral plane. The path is symmetrical to equally subsample the space in front of the user. **c.** Example session showing the lateral (top) and upward (middle) position of the target, fingertip, and gaze point for a right-hand trial. Bottom: Euclidean distance from the fingertip to the target (target error). **d, e.** 5-cm binned positional data comparing the target position to the fingertip position in the lateral (top) and upward (middle) dimensions for left (**d**) and right (**e**) hand trials. Insets show binned positional data comparing the gaze point position to the target position. Bottom plots show target error as a function of target speed.

The target in this task was a 4-cm sphere that moved in a pseudo-random 2D path at 75% of the user’s reach. Once the user has touched the sphere with their index finger for 1 second it starts moving along this predefined path. The upward and lateral components of the path were the sum of sine waves which were designed to subsample each quadrant in front of the user equally (Figure 6b, see Methods). From the user’s perspective the path moves randomly, meaning it could be repeatedly presented from session to session without any ability to predict the motion, yielding an untrained estimate of motor function.

Fingertip and gaze point data were extracted during the tracking interval and analyzed in the lateral and upward directions (Figure 6c). We computed the Euclidean distance between the fingertip and target at each point – target error – to estimate the user’s ability to track the target. To quantify the tracking in this task we binned the positional data of the target in 5-cm intervals and calculated the position of the fingertip and gaze point in each of these bins (Figure 6d,e). The slope of a linear regression of binned data is a proxy for the user’s accuracy to track the target, with slopes closer to 1 indicating better tracking. Fingertip data was a close match to target position across the bins (Right hand: linear regression of lateral position: slope = 0.723, p-value < 1 x 10^-15^; linear regression of upward position: slope = 0.790, p-value < 1 x 10^-15^; Left hand: linear regression of lateral position: slope = 0.715, p-value < 1 x 10^-15^; linear regression of upward position: slope = 0.771, p-value < 1 x 10^-15^), however gaze point data was less of a close fit, especially in the lateral dimension (Right hand trial: linear regression of lateral position: slope = 0.165, p-value < 1 x 10^-15^; linear regression of upward position: slope = 0.388, p-value < 1 x 10^-15^; Left hand trial: linear regression of lateral position: slope = 0.147, p-value < 1 x 10^-15^; linear regression of upward position: slope = 0.382, p-value < 1 x 10^-15^). This is possibly the result of low framerates on HoloLens cameras responsible for eye tracking. Further analysis of gaze point data in this task on movement disorder patients would help us understand whether this is a useful task for tracking eye movements. Last, we also measure target error of the fingertip as a function of speed to assess the user’s responsivity to the target (Figure 6d,e).

### Vocal assessment task

Last, to demonstrate the scope of behavioral tracking that could be accomplished with mixed-reality headsets in the clinic, we designed a task to assess the vocal qualities of a user while reading phrases aloud. Vocal changes are pathognomonic of many neurological disorders including Parkinson’s disease^20^, progressive supranuclear palsy^21^, and cerebellar damage^19^, thus quantification of the voice along with the other tasks above could add depth to the diagnostic capabilities of physicians. To quantify features of the user’s voice, we designed a task where users would be presented with written phrases that they were instructed to read aloud. These phrases were a combination of natural sentences and phrases designed to test the user’s ability to say lingual, labial, and guttural sounds.

At each presentation of a phrase, a 5 second audio clip was recorded for offline processing. We used the open-source package Parselmouth, which ports functionality from the commonly used speech processing software Praat into Python^22,23^. In each audio clip we analyzed features of the vocal frequency, amplitude, and volume (Figure 7a). We used Parselmouth to extract the speaking time, intensity of the voice in decibels, vocal pitch (mean frequency of the fundamental formant), shimmer (amplitude variation of the sound wave), and the local jitter (the frequency variation from cycle to cycle of F0)^24^. The mean of these features across all spoken phrases was used to compute a vocal fingerprint of the user (Figure 7b; mean speaking time = 0.746 s, mean intensity = 56.28 dB, mean F0 pitch = 101.1 Hz, mean local shimmer = 0.133 dB, mean local jitter = 0.0352 Hz). This fingerprint could be used to assess longitudinal changes to the user’s voice to monitor disease progression or treatment efficacy.

**Figure 7.**
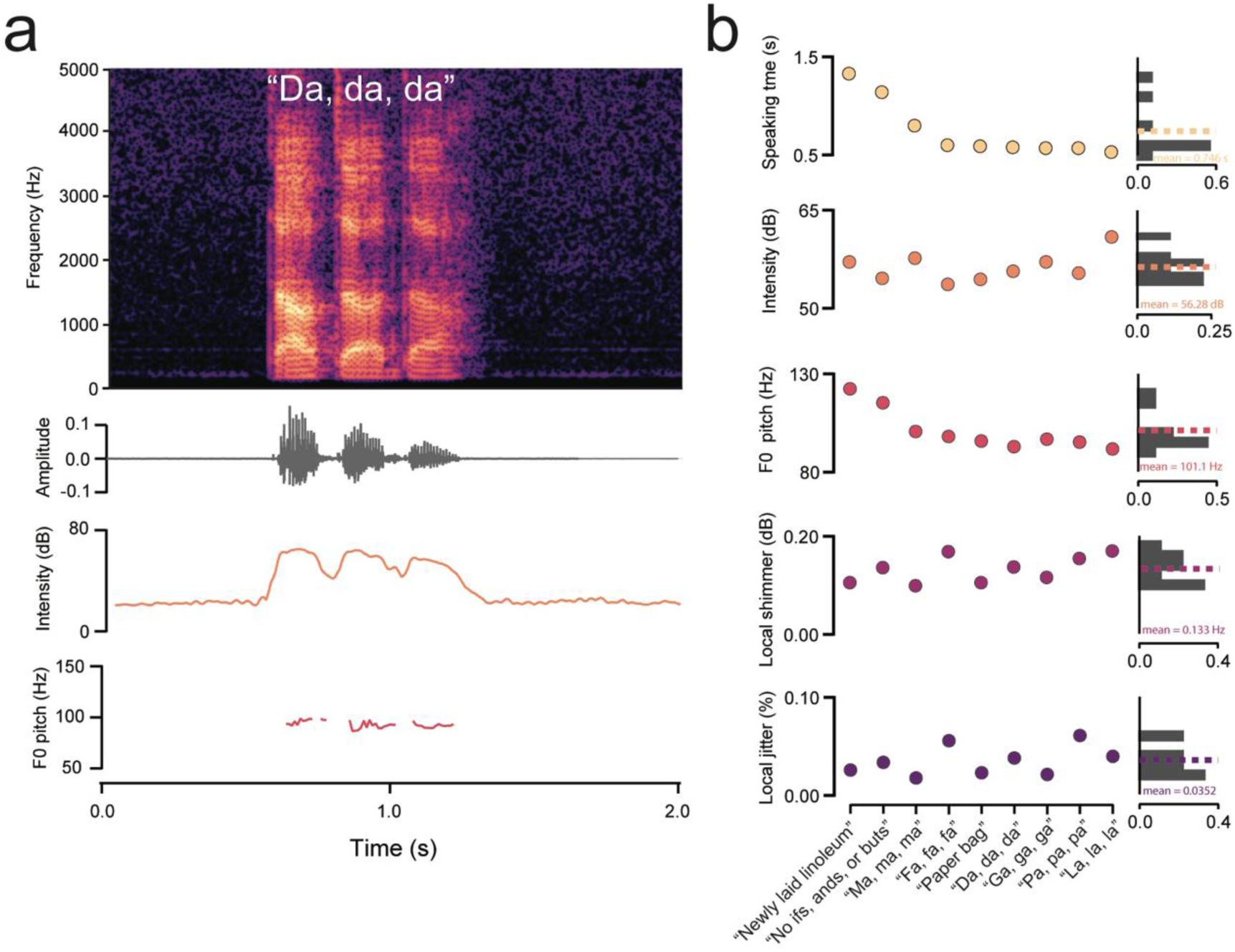
Quantification of vocal features evoked with the vocal assessment task. **a.** Top: frequency spectrogram of a user saying the phrase “Da, da, da.” Second from top: Amplitude of the phrase over time. Third from top: Intensity of the phrase in decibels. Bottom: Pitch of the fundamental formant of the phrase plotted during periods of detected speech. **b.** Left: Quantification of the speaking time, intensity, F0 pitch, local shimmer, and local jitter of 9 phrases ordered by length of speaking time. Right: Histograms of vocal features with mean (dashed line).

## Discussion

Here we show proof of concept of an application on the Microsoft HoloLens 2 mixed-reality headset that can be administered with little to no input from hospital staff while capturing thorough, high-fidelity behavioral data. As a starting point, we designed 6 tasks that replicate aspects of the neurologic examination and show the capacity of mixed reality to probe upper extremity movement, eye tracking, and vocal analysis.

Patients with movement disorders often face lengthy wait times to see movement disorder specialists that can diagnose and monitor progression of disease. The American Academy of Neurology Workforce Survey suggests that wait times to see neurologists are increasing and that the neurologist shortfall for the US will reach 19% by 2025^25^. These trends are worse in regions deemed “neurology deserts” that are particularly lacking in neurologic care^26^. An application, like the one described in this paper, could help solve this issue in several ways. First, because headsets like the HoloLens 2 are commercially available and mass-produced, they can be distributed to patients themselves, health fairs, pharmacies, primary care offices, or general neurology offices. Widespread access could enable low-cost screening for movement disorders allowing patients to be identified and escalated to a specialist for diagnosis. Modeling suggests that community screening for Parkinson’s disease using low-cost technology can be a cost-effective way to increase quality-adjusted life years if deployed in public places such as health fairs and pharmacies^27^. Second, for patients who already have a diagnosis, mixed-reality headsets could provide more frequent monitoring of disease progression over time and in response to treatment. Rather than assessing disease progression and adjusting medications at follow up appointments every 3-6 months, patients could use a headset in between appointments and then bring their results to their follow up appointment to give the neurologist a more holistic picture their day-to-day variability. This could decrease the frequency of in person follow up appointments and improve the quality of telehealth assessments, which would be particularly beneficial in rural areas without access to movement disorder neurologists.

Consistent presentation of stimuli and objective, high-fidelity quantification of behavior will decrease measurement variability, which could be useful for assessing efficacy of interventions. Lower assay variability allows for better discrimination of positive and negative effects and smaller samples sizes. Additionally, the objective nature of the mixed-reality headset enforces blinding and removes the potential for subjective bias from the clinician rating the exam. This could allow for more robust and less expensive clinical trials to test new drug candidates and therapeutic interventions for movement disorders.

Mixed reality headsets and holograms also allow for new tasks that are not yet part of the standard neurologic exam and might shed light on previously unappreciated neurologic deficits. Such tasks could include puzzles that require manipulation of 3D objects, navigation tasks, or tests of spatial memory. We speculate that there are likely unrecognized subsets of neurologic diseases that could be revealed by novel tasks and deep analytic algorithms that can cluster patient data and point neurologists towards outliers and novel phenotypes.

Mixed-reality tasks could also be beneficial for patients with deep brain stimulators (DBS) that require periodic reprogramming. DBS is currently used in patients with Parkinson’s disease, essential tremor, and dystonias. Occasionally, DBS probes must be reprogrammed to ensure they are targeting the correct areas and managing symptoms appropriately. More than a third of PD patients report difficulty getting to a clinic^26^ and consequently, tele-programming has started to emerge^28^. Mixed-reality headsets could allow clinicians to optimize DBS programming both in person and over telehealth through the use of short, quantifiable tasks that enable rapid iteration and objective comparison between providers over spans of months and years.

In future work, our paradigm will be applied to patients with movement disorders such as Parkinson’s disease to test our ability to discriminate between patients with and without disease. Mixed-reality headsets like the HoloLens 2 could serve as the black leather physician bags of the 21st century neurologist; containing tools to probe and quantify movement, vision, and vocal production, enabling better access to care, and allowing for the discovery of new phenotypes and disease subtypes.

## Methods

### HoloLens application design

Applications were built using Unity software (2020.3.21f1) and uploaded to the Microsoft HoloLens 2 headset using Visual Studio (2019). In each application, behavioral data of interest (hand, eye, voice, events) was written to a text file that was then exported to a computer for post-processing. Kinematic data were written to the text files at 60 Hz. Event times were written to separate text files following important events during each task including task start time, task end time, countdown start times, countdown stop times, target touch times, and target move times. Hand data consisted of positional data in 3 dimensions from all tracked points on the right and left hand: the wrist, palm, carpometacarpal joints of each finger and thumb, metacarpophalangeal joints of each finger and thumb, proximal interphalangeal joint of each finger and thumb, distal interphalangeal joint of each finger, and fingertips of each finger and thumb for a total of 26 tracked joints. Gaze origin and gaze ray were written to capture eye movements in three directions. Vocal data were saved as .wav audio files to the headset.

### Post-processing of HoloLens data

HoloLens positional data was saved as distance in meters in the lateral (x), upward (y), and outward (z) directions for postprocessing offline. Data were resampled at 60 Hz then filtered with a 2^nd^ order 10 Hz lowpass Butterworth filter forwards then backwards to avoid phase shift^29^. Data were clipped into periods of interest by using the timestamps of event data written to the headset.

To analyze tremor in the tremor assessment task, we computed the average speed and acceleration during the countdown period by calculating the frame-to-frame changes in position (speed) or speed (acceleration) and dividing by the sampling rate. The power spectrum of each point on the hand was computed as described above. To analyze tapping movement, we computed tap amplitude and tap times as described above. To analyze changes in tap time and tap amplitude across the trial, we subjected these variables to a linear regression, with the slope indicating the change in tap time or amplitude across the trial. For pronation-supination analysis, we first applied a rotation matrix to align the hand to the Y-axis at each frame of saved data. Specifically, we aligned a hand vector (the vector projecting from the wrist to the middle finger MCP) to the Y-axis by solving for this rotation matrix analytically. Once the hand was aligned, we measured the angle of a vector projecting from the middle finger MCP to index finger MCP, around the Y-axis to yield a turn angle. We normalize this angle such that 0 degrees was the median value of each pronation-supination trial. Turn amplitude and time were analyzed as described for tapping above. To analyze center out data, we isolated individual trials by extracting index fingertip positions between “target move” event data timestamps. We computed the angle of target motion, such that trials can be grouped by movement direction. Speed and tortuosity were computed as described above. To analyze random tracking, we measured index fingertip position and target position binned in 5-cm intervals in target position location. Target error was computed as the Euclidean distance between index fingertip position and target position. To calculate “gaze point” location in all tasks, we found the shortest line between the target and the gaze ray and found where this line intersected the gaze ray. This point was used as a proxy to understand where the user was looking in 3D space. Vocal analysis was performed using Parselmouth^23^.

### Random path construction

The pseudo-random path that users followed in the random tracking task was constructed by summing 100 sine waves with random frequencies between 1 and 10 Hz and offset by random phase shifts from 0 to 1 second in 1/60 s intervals. Here Hz refers to the length of the trial (*i.e.,* 1 Hz = 1 cycle per trial). This was performed 2 times to construct a waveform that described target motion in the lateral and upward dimensions. The result was a random trajectory in lateral and upward space. The magnitude of these waves was normalized so that the maximum value in each dimension was 30 cm.

### Metronome stimulus

We used an online metronome to deliver an auditory tone to provide an external stimulus to drive movements. For tapping and pronation-supination experiments users made movements at 80, 120, and 160 bpm (0.75, 0.5, and 0.375 second interbeat interval, respectively). For tremor experiments the hands were moved up and down in the vertical dimension at 60, 120, 180, and 240 bpm (1-4 Hz).

## Data Availability

All data produced in the present study are available upon reasonable request to the authors

---

**Figure S1.**
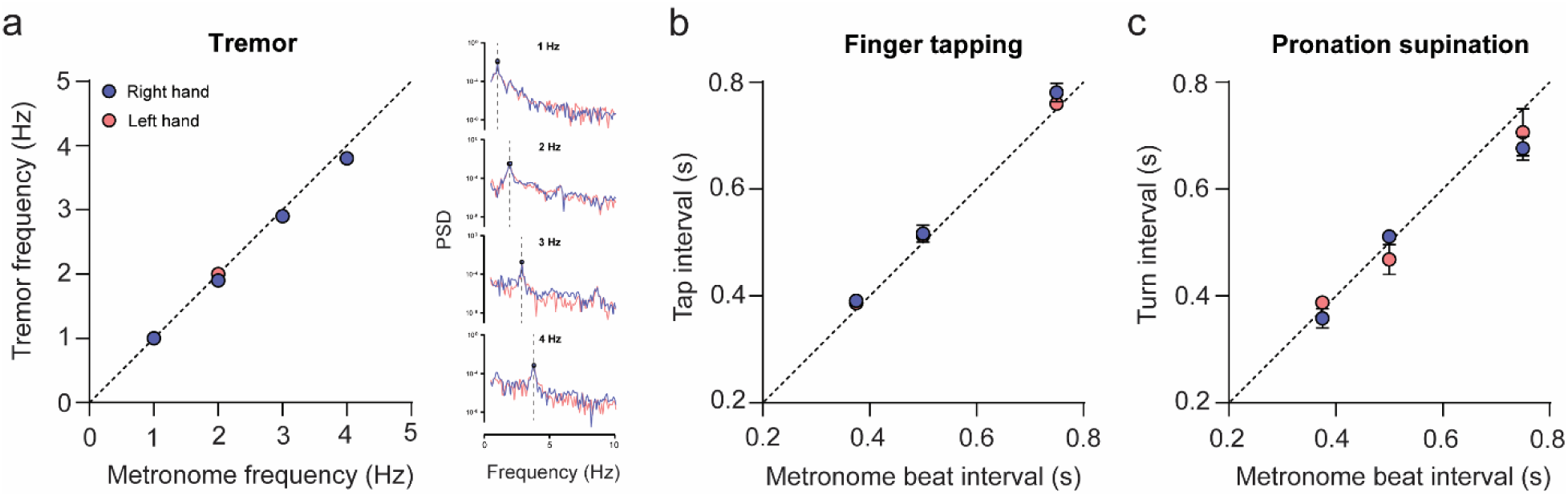
Hand movements match external driving stimuli. **a.** Left: The frequency of positional tremor closely matched the metronome driving frequency at 1-4 Hz. Right: Power spectral density of upward position of the palm of the right (red) and left (blue) hands. The peak frequency is denoted with a point and vertical dashed line. Note: Only Upward position was analyzed because the tremor was simulated by moving hands up and down at the rate of the metronome. **b.** Kinematic tracking of finger tap times closely matched the metronome beat interval across a range of frequencies for both the right (blue) and left (pink) hands. **c.** Same as **b**, but for pronation-supination movements.

## References

1. Fahn, S., Marsden, C., Calne, D., Goldstein, M. & Editors. Members of the UPDRS Development Committee. in Recent Developments in Parkinson’s Disease, Vol 2. 153–163, 293–304 (Macmillan Health Care Information, 1987).

2. Siesling, S., Zwinderman, A. H., van Vugt, J. P., Kieburtz, K. & Roos, R. A. A shortened version of the motor section of the Unified Huntington’s Disease Rating Scale. Mov Disord 12, 229–234 (1997).

3. Subramony, S. H. SARA—a new clinical scale for the assessment and rating of ataxia. Nat Clin Pract Neurol 3, 136–137 (2007).

4. Weyer, A. et al. Reliability and validity of the scale for the assessment and rating of ataxia: A study in 64 ataxia patients. Movement Disorders 22, 1633–1637 (2007).

5. Sato, K. et al. Reliability of the Japanese version of the scale for the assessment and rating of ataxia (SARA). Brain and Nerve 61, 591–595 (2009).

6. Gavgani, A. M., Nesbitt, K. V., Blackmore, K. L. & Nalivaiko, E. Profiling subjective symptoms and autonomic changes associated with cybersickness. Auton Neurosci 203, 41–50 (2017).

7. Duzmanska, N., Strojny, P. & Strojny, A. Can simulator sickness be avoided? A review on temporal aspects of simulator sickness. Front Psychol 9, (2018).

8. Guna, J. et al. Influence of video content type on users’ virtual reality sickness perception and physiological response. Future Generation Computer Systems 91, 263–276 (2019).

9. Lawson, B. D. & Stanney, K. M. Editorial: Cybersickness in Virtual Reality and Augmented Reality. Front Virtual Real 2, (2021).

10. Sternberg, E., Alcalay, R., Levy, O. & Louis, E. Postural and Intention Tremors: A Detailed Clinical Study of Essential Tremor vs. Parkinson’s Disease. Front Neurol 4, (2013).

11. Charles, P. D., Esper, G., Davis, T. L., Maciunas, R. J. & Robertson, D. Classification of tremor and update on treatment. Am Fam Physician 59 6, 1565–72 (1999).

12. Simonet, C. et al. Slow Motion Analysis of Repetitive Tapping (SMART) Test: Measuring Bradykinesia in Recently Diagnosed Parkinson’s Disease and Idiopathic Anosmia. J Parkinsons Dis 11, 1901–1915 (2021).

13. Djurić-Jovičić, M. et al. Finger tapping analysis in patients with Parkinson&#x2019;s disease and atypical parkinsonism. Journal of Clinical Neuroscience 30, 49–55 (2016).

14. Gulde, P., Vojta, H., Hermsdörfer, J. & Rieckmann, P. State and trait of finger tapping performance in multiple sclerosis. Sci Rep 11, 17095 (2021).

15. Shirani, A., Newton, B. D. & Okuda, D. T. Finger tapping impairments are highly sensitive for evaluating upper motor neuron lesions. BMC Neurol 17, (2017).

16. Roalf, D. R. et al. Quantitative assessment of finger tapping characteristics in mild cognitive impairment, Alzheimer’s disease, and Parkinson’s disease. J Neurol 265, 1365– 1375 (2018).

17. Agostino, R., Berardelli, A., Currà, A., Accornero, N. & Manfredi, M. Clinical impairment of sequential finger movements in Parkinson’s disease. Movement Disorders 13, 418–421 (1998).

18. Montgomery, E. B., Nuessen, J. & Gorman, D. S. Reaction time and movement velocity abnormalities in Parkinson’s disease under different task conditions. Neurology 41, 1476– 1481 (1991).

19. Goodlett, C. R. & Mittleman, G. Chapter 9 - The Cerebellum. in Conn’s Translational Neuroscience (ed. Conn, P. M.) 191–212 (Academic Press, 2017). 10.1016/B978-0-12-802381-5.00016-6.

20. J. Holmes, R., M. Oates, J., J. Phyland, D. & J. Hughes, A. Voice characteristics in the progression of Parkinson’s disease. Int J Lang Commun Disord 35, 407–418 (2000).

21. Skodda, S., Visser, W. & Schlegel, U. Acoustical Analysis of Speech in Progressive Supranuclear Palsy. Journal of Voice 25, 725–731 (2011).

22. Boersma, P. Praat, a system for doing phonetics by computer. in (2002).

23. Jadoul, Y., Thompson, B. & de Boer, B. Introducing Parselmouth: A Python interface to Praat. J Phon 71, 1–15 (2018).

24. Teixeira, J. P., Oliveira, C. & Lopes, C. Vocal Acoustic Analysis – Jitter, Shimmer and HNR Parameters. Procedia Technology 9, 1112–1122 (2013).

25. Freeman, W. D., Vatz, K. A., Griggs, R. C. & Pedley, T. The Workforce Task Force Report Clinical implications for neurology. www.neurology.org (2013).

26. Majersik, J. J. et al. A Shortage of Neurologists – We Must Act Now. Neurology 96, 1122 (2021).

27. Muñoz, D. A., Kilinc, M. S., Nembhard, H. B., Tucker, C. & Huang, X. Evaluating the cost-effectiveness of an early detection of Parkinson’s disease through innovative technology. Engineering Economist 62, 180–196 (2017).

28. Esper, C. D. et al. Necessity and feasibility of remote tele-programming of deep brain stimulation systems in Parkinson’s disease. Parkinsonism Relat Disord 96, 38–42 (2022).

29. Yu, B., Gabriel, D., Noble, L. & An, K. N. Estimate of the optimum cut-off frequency for the Butterworth low-pass digital filter. J Appl Biomech 15, 318–329 (1999).

